# Digital Detection of Prolonged Grief Disorder (PGD) in Vietnamese Bereaved: Protocol for an Experience Sampling Study

**DOI:** 10.1101/2025.10.22.25338429

**Authors:** Huy Hoang Le, Hung Nguyen, Nhien Du, Cat Pham, Trang Le, Hoang-Minh Dang, Clare Killikelly

## Abstract

**Purpose:** Prolonged Grief Disorder (PGD), recently included in the DSM-5-TR and ICD-11, involves a strong yearning for the deceased, fixation on death, denial, avoidance, negative emotions, and disengagement from daily life, lasting at least six months. However, most studies focus on Western settings, leading to underdiagnosis, inadequate care, and a growing mental health burden in Viet Nam.

**Design:** Experience sampling method (ESM) can provide detailed, real-time data on daily symptoms due to the wave-like nature of grief and its psychological sequelae. 100 Vietnamese bereaved individuals who lost a loved one in the last 6 months will complete a 2-week ESM assessment app (mPath) every 3 months for 6 months. The app sends daily reminders to complete a questionnaire about PGD symptoms and daily life context. Participants will complete a baseline mental health assessment before each ESM period.

**Findings:** Data collection for ESM assessments began in June 2025 and will end in December 2025. As of September 2025, 91 participants have been enrolled. A mix of longitudinal and fine-grained data will reveal daily fluctuations and chronicity of PGD symptoms, identify prodromal symptoms predicting onset and severity, unique core symptoms to the Vietnamese bereaved, and factors predicting onset.

**Originality:** This study will be the first to not only digitally screen for mental health distress in Viet Nam, but also the first to study the newly defined condition PGD in Viet Nam.

## Introduction

### Background

Worldwide, rates of mental health disorders have surged since the onset of the COVID-19 pandemic in 2019. The WHO estimates a 25% increase in depressive and anxiety disorders (Santomauro *et al*., 2021). In Viet Nam, studies on mental health have estimated the prevalence of post-traumatic stress disorder (PTSD), depression, and anxiety during the pandemic at 22.9%, 11.2%, and 17.4%, respectively (Nguyen *et al*., 2023). Bui *et al*. (2024) estimated that the prevalence of depressive and anxiety symptoms has risen to 51.92% and 56.75%, with notably high stress symptom prevalence at 43.64%. Post-COVID-19, researchers and clinicians have been preparing for a "shadow pandemic" of mental health disorders related to bereavement (Eisma *et al*., 2020). Alongside the rapid and traumatic nature of COVID-19-related deaths, social restrictions and lockdowns have exposed many people to complex and isolated grieving situations (Sun *et al*., 2020).

A large-scale study confirmed that COVID-19 losses led to elevated rates of acute grief, prolonged grief, PTSD, and other stress-related symptoms (Eisma *et al*., 2021). In East and Southeast Asia, such losses are often considered “bad deaths” due to their unnatural cause and the social restrictions that disrupt mourning rituals (Hinton *et al*., 2013). Families experiencing “bad deaths” face heightened risks for pathological prolonged grief, exacerbated by limited access to mental health care during the pandemic (Baumgart *et al*., 2021; Carr *et al*., 2020). For each death, an estimated nine family members experience emotional distress, with 60% recovering naturally and 40% at risk for mental health issues, including about 10% with severe, persistent grief (Bonanno *et al*., 2011; Pearce *et al*., 2021; Szuhany *et al*., 2021). In Viet Nam, over 43,000 COVID-19 deaths as of April 28, 2022, likely resulted in approximately 388,000 mental health impacts, with more than 155,000 individuals needing grief support. Prolonged Grief Disorder (PGD), a newly recognized diagnosis, has significant knowledge gaps. Clinicians require clearer methods to differentiate PGD from normal grief (Keeley *et al*., 2016), and targeted interventions must be developed (Shear *et al*., 2016). While early diagnosis benefits depression and PTSD, premature grief intervention can be harmful (Prigerson *et al*., 2000). Early PGD symptoms can predict later PTSD (Djelantik *et al*., 2017; O’Connor *et al*., 2015) and depression (Boelen & Lenferink *et al*., 2020), though results vary when PGD, PTSD, and MDD are assessed together—e.g., PTSD, not PGD, predicted later depression after traumatic loss (Boelen & Boelen & Lenferink et al., 2020). Bereavement can also lead to adjustment disorder, mood, and anxiety disorders (Boelen *et al*., 2016; Horowitz *et al*., 1997; Jordan & Litz, 2014). Around 10% of bereaved individuals develop PGD (Lundorff et al., 2017), 12–16% PTSD (O’Connor *et al*., 2010), and 19% depression (Blanner Kristiansen *et al*., 2019); among those with PGD, 40% also have PTSD and 63% depression (Komischke-Konnerup *et al*., 2021). Accurate, timely diagnosis is essential, as PGD responds poorly to treatments for MDD or PTSD, and vice versa (Shear *et al*., 2016).

Notably, extensive epidemiological research on PGD has predominantly been conducted within Western contexts, primarily involving older White women (Maciejewsky *et al*., 2016; Prigerson *et al*., 2021). Methodological imprecision and a lack of consideration for sociocultural contexts surrounding mourning and bereavement have hindered the consistent generation of global findings in multicultural comparative grief research (Stelzer *et al*., 2020; Silverman *et al*., 2021). Consequently, the prevalence rates of PGD vary significantly worldwide. Recent meta-analyses reported a global prevalence rate of 9.8%, with middle-income East Asian countries reporting lower rates (8.9% - 9.2%) compared to Western countries (10.2%) (Lundorff *et al*., 2017; Le, 2024). However, these regional averages are largely based on data from China and Japan, which may not accurately reflect the experiences of bereaved individuals in other Asian nations. Applying findings from a few countries to the entire region risks overlooking important contextual factors. Viet Nam, in particular, has received little attention in PGD research, despite its unique sociocultural landscape and the growing burden of grief-related mental health issues. This gap underscores the urgent need for studies investigating PGD development and presentation among the Vietnamese population.

Given the fluctuating nature of grief reactions, app-based experience sampling method (ESM) can capture real-time PGD reactions in daily life (Lenferink *et al*., 2023). Rooted in ecological psychology, ESM emphasizes understanding symptoms in real-world contexts (Myin-Germeys *et al*., 2018). Mobile apps are more accessible than paper-based tools or dedicated devices (Granholm *et al*., 2012) and reduce biases tied to recall, reliability, and averaging in clinical monitoring (Palmier-Claus *et al*., 2013). ESM data can identify triggers, relapse signs, treatment effects, and early symptom changes (Gumley *et al*., 2022). It has been applied to track symptom variability and trajectories in depression, PTSD, PGD, and schizophrenia (Depp *et al*., 2016; Kimhy *et al*., 2014; Lenferink *et al*., 2025), predict depression (Fried *et al*., 2024; Minaeva *et al*., 2020), and flag suicide risk or deterioration in psychosis (Lewis *et al*., 2020). Recent studies found ESM to be acceptable and feasible for recently bereaved individuals (Aeschlimann *et al*., 2024; Lenferink *et al*., 2022). In Viet Nam, it was successfully implemented with a 61.8% compliance rate (Trang *et al*., 2022).

This research endeavor contributes to the ongoing global efforts in transforming general healthcare and mental healthcare practices via digitalization. Yet, current usages are limited to digital medical records and digital prescriptions (Orsolini *et al*., 2025). The recent global survey by Orsolini and colleagues (2025) also suggested that the widespread availability of mobile applications in general and mental health care did not translate into a high level of usage, especially for screening and diagnosis of mental illness. In Viet Nam, the country’s broader push toward digital health transformation (Dang *et al*., 2021) and positive attitudes toward healthcare digitalization among healthcare providers (Pham *et al*., 2025) have shown progress in digital healthcare, including machine learning for carcinogenic risk detection (Le *et al*., 2024) and COVID-19 surveillance and contact tracing (Bui *et al*., 2021). It may, however, be noted that digital mental healthcare practices in Viet Nam follow global trends. That is, to the best of our knowledge, there has only been 1 app-based mental health intervention adapted for use in Viet Nam (Chau *et al*., 2023; 2025; Murphy *et al*., 2020), and no report has been found for the usage of digital screening and diagnosis of mental illness in Viet Nam.

### Objective

This field is currently at a critical turning point, where clinicians and researchers are urgently called upon to provide treatment and support for those bereaved under stressful circumstances before, during, and after the pandemic, while also adapting to the introduction of a newly defined grief-related disorder (Killikelly *et al*., 2021). For this reason, objective and culturally sensitive research is required to predict trajectories of grief following bereavement to address the increasing demand for mental health care among the Vietnamese community. This paper presents the protocol for a study that aims to address the following questions:

1. When do PGD symptoms become clinically significant? What is the temporal relationship between the onset of PGD symptoms and the diagnosis of PGD disorder?
2. Can we identify early signs (prodromal symptoms) that predict the onset and clinical severity of the disorder?
3. Can we identify novel factors related to disorder/recovery status that emerge during the disorder’s progression?
4. How do symptoms progress over time (symptom variability and chronicity)? Is there variability between individuals and within individuals regarding core PGD symptoms and comorbid conditions?

## Methods

### Participants

100 adult participants will be enrolled in the study. Eligibility for this study includes participants who have lost a loved one (e.g., family members, spouse, close friends, etc.) 3-6 months, are currently not seeking any mental health services to cope with the loss, and have never received a formal psychiatric diagnosis will be recruited via online channels (e.g., Facebook groups and LinkedIn).

### Ethical considerations and declarations

Ethical approval was obtained from the ethics committee of Viet Nam National University – University of Education (No. 25.05/HDDD-DHGD). Informed consent will be collected during the virtual baseline assessment. Participants can withdraw at any time and receive 1.2 million Vietnamese Dong for participation. Patient identifiers will be anonymized, and new identifiers will replace names. Data will be securely stored in password-protected files and publicly available for reanalysis with meticulous deidentification. Identifying information won’t be disclosed for publications or presentations. Bank account information of eligible participants will be stored for compensation. All research data will be two-factor password-protected and stored on secure servers of the University of Zurich.

### Materials

In this study, we have decided to utilize the digital app m-Path (www.mPath.io/dashboard), an online platform that provides an easy-to-use and highly tailorable framework, for our smartphone-based ESM. The app was translated into Vietnamese by the first author (HL). Baseline assessment was conducted virtually via LimeSurvey (http://limesurvey.org).

### Measures

Depressive symptoms were measured with the Patient Health Questionnaire-9 (PHQ-9), validated in Vietnamese adults and clinical populations (Nguyen *et al*., 2020; Phi *et al*., 2023). Generalized anxiety was assessed with the Generalized Anxiety Disorder-7 (GAD-7), validated in Vietnamese clinical settings (Nguyen, 2021). Somatic symptoms were measured with the Somatic Symptom Scale–8 (SSS-8), used in Viet Nam for behavioral health integration research (Wu *et al*., 2022). Other measures included the WHO Wellbeing Index (WHO-5), International Trauma Questionnaire (ITQ) (Cloitre *et al*., 2018), International Prolonged Grief Disorder Scale (IPGDS) (Killikelly *et al*., 2020), Oxford Grief Social Disconnection Scale (OG-SD), Depressive and Anxious Avoidance in Prolonged Grief Questionnaire (DAAPGQ), and MyGrief items (O’Connor et al., in preparation), and a self-developed grief monitoring scale.

The daily ESM survey included 3 IPGDS items, 9 MyGrief items (modified for ESM), 3 daily life context questions (e.g., "What were you just doing?"; "Where are you right now?"; "Who are you with right now?"), and 3 enjoyment questions related to activity, place, and person (e.g., “How much do you enjoy this activity?”; “How much do you enjoy being here?”; “How much do you enjoy being around this person?”).

### Procedures

For questionnaires not available in Vietnamese, the research team will use Fenn et al. (2020)’s standard translation process to ensure quality. At least two independent translators will translate forward: one fluent in Vietnamese but not psychology, and the other fluent in Vietnamese with psychological expertise. At least two independent translators will back-translate, without involvement in the forward translation. Finally, a panel of experts will review and compare the translations for accuracy and appropriateness.

In May 2025, a cognitive validity interview process was conducted to identify misalignment between participants’ interpretations and the researcher’s. The process evaluated the clarity and language adaptation of the ESM items after translation, ensuring they were understandable and culturally appropriate for the Vietnamese context. Participants imagined themselves as research participants and answered questions based on Peterson and colleagues’ (2017) cognitive operations. They clarified difficult words, suggested alternative words, and identified words for change. After the comprehension review, participants compared the items, identifying any easy, difficult, or sensitive items for reformulation. The research team revised the questions to improve comprehension and cultural relevance.

To schedule daily assessments, the researcher (HL) logged into m-Path’s online dashboard at www.m-Path.io/dashboard with an existing account to create a “Protocol”, an ESM template consisting of the previously validated questions, to apply to future participants. The Protocol for this study was scheduled to send participants 5 reminders daily (1 in the morning, 1 at noon, 1 in the afternoon, and 2 in the evening) to fill out the questionnaire. See Fig. 1 for our m-Path workflow.

**Fig 1.**
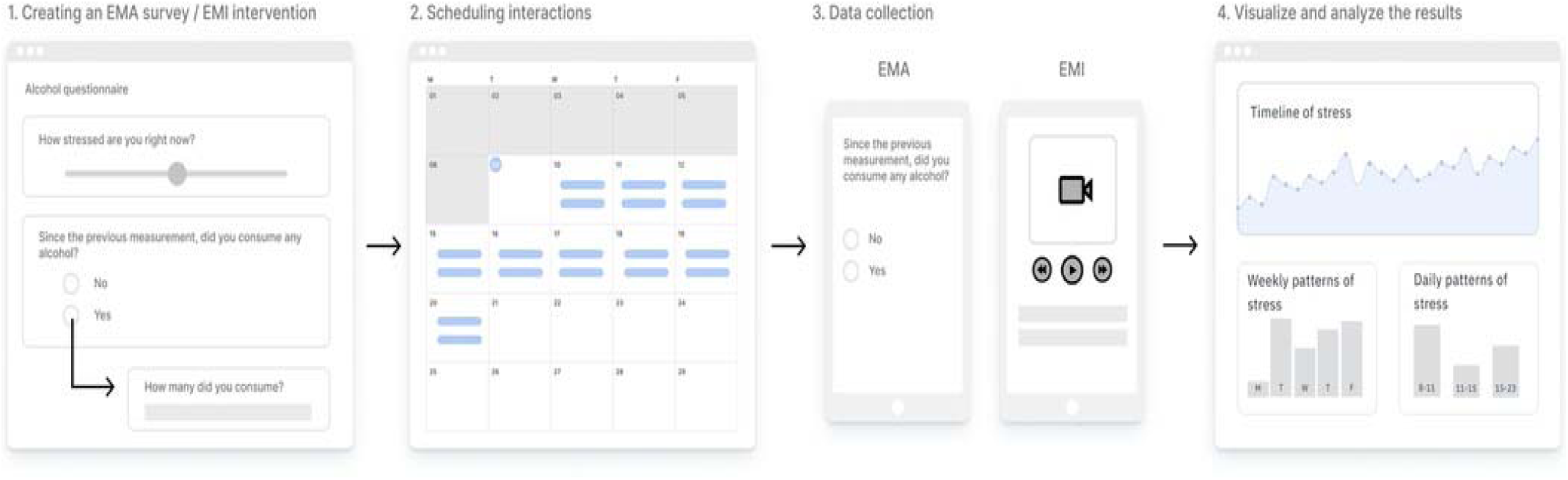
The m-Path workflow. First, researchers create and fine-tune the ESM survey in the interaction editor (Panel 1). Next, these ESM interactions are scheduled in the calendar view (Panel 2). Third, participants receive notifications on their personal smartphones to interact with the ESM content (Panel 3). Finally, researchers can analyze incoming results of a single participant in real-time via modifiable graphs and charts in the online dashboard (Panel 4). Reproduced from Mestdagh, Verdonck, Piot, Niemeijer, Kilani, Tuerlinckx, Kuppens, and Dejonckheere, 2023, Frontiers in Digital Health, 5:1182175 [62] under the terms of the Creative Commons Attribution License (CC BY).*Data collection:*

Step 1: Participants provide personal details (name, email, birthdate, relationship to the deceased, date of passing). Eligible participants receive an email to set up a call showing how to download and use the mPath app. They complete a 5-minute trial run and receive a follow-up email with detailed instructions. Participants are invited to participate in the baseline assessment and sign an online consent form.

Step 2: Baseline Assessment (PreESM): The assessment will take about 1 hour. Before each ESM experience sampling survey, participants will be required to complete an online mental health status assessment on the Lime Survey survey tool. Participants will complete this assessment two more times, after 3 months and 6 months.

Step 3: ESM Data Collection: Participants will fill out an integrated questionnaire on the m-Path about prolonged grief disorder (PGD) symptoms and their daily life context 5 times daily for over two weeks. They’ll complete the mobile app sampling 3 times, each lasting 2 weeks with 3-month intervals, over a 6-month period.

### Data analysis plan

Psychometric property: Cronbach’s alpha coefficient and McDonald’s Omega will be used to measure the reliability of the Vietnamese version of the IPGDS. Confirmatory Factor Analysis (CFA) will be conducted on the translation to examine whether it maintains the same structure as the original version.

Treatment of missing data: Patterns of missing data will be visualized using the “naniar” tool in R, and missing completely at random (MCAR) will be assessed via Little’s MCAR test. If missingness is identified as MCAR, multiple imputation will be implemented. Otherwise, missing at random (MAR) is assumed, and full information maximum likelihood (FIML) will be applied for handling missing data. Additionally, sensitivity analysis is conducted to account for possible missing not at random (MNAR), to which pattern-mixture models will be applied (Staudit et al., 2022).

Clinical PGD onset will be assessed via survival and logistic regression analyses using PGD symptoms across 3 baseline timepoints. Cross-lagged panel model will be used to assess bidirectional temporal relationships between symptom onsets and PGD severity. Analysis of individual variability will explore predictive models; for example, the emergence of prodromal symptoms can be compared to PGD deterioration at the next assessment point to test the predictive validity of disorder development (see Gaebel & Riesbeck, 2014). Diagnostic item-level analysis of PGD can also be used to detect increases in specific items on the ESM (weighted moving average with an exponential function, Smit & Snippe, 2022), such as increased longing for the deceased, to check if this increase occurs in individuals who later develop psychological disorders. Following previous ESM research on grief (Lenferink et al., 2024), Root Mean Square of Successive Differences (RMSSD) will reflect day-level variability on ESM grief items. RMSSD comprises two components: variability (within-person variance, or dispersion) and temporal dependency (autocorrelation). Higher WPV indicates greater symptom fluctuation, while higher autocorrelation signifies greater resistance to change. Larger RMSSD values indicate greater fluctuations.

We will employ latent class growth modeling (LCGM) to identify distinct longitudinal patterns of Prolonged Grief Disorder (PGD) symptoms. These models capture heterogeneity in individual change trajectories, distinguishing between subgroups with different symptom persistence, escalation, or recovery courses. We will determine the optimal number of trajectories based on: (1) lower AIC and SSA-BIC; (2) higher entropy R^2^ (> .80 acceptable, 0.60-0.80 moderate-good fit); and (3) significant p-values (< .050) for VLMR-LRt, LMR-LRt, and BLRt, indicating a better fit than a model with one fewer class (Nylund et al., 2007). After determining the best-fitting model, we will apply a three-step approach to preserve class assignments and examine predictors of trajectory membership using multinomial logistic regression, with baseline PGD severity, within-person daily PGD fluctuations, demographic characteristics, bereavement-related factors, and prior health status as covariates. In addition, binary logistic regression will be conducted to evaluate the role of these predictors in the development of clinically significant PGD or in achieving sustained recovery, providing complementary insights into both classification and mechanisms of progression or improvement.

## Results

The cognitive validity interview for the ESM questionnaire was completed in May 2025. Researchers interviewed 13 adults (Mage = 28.46, 6 female, 7 male) who had lost a loved one and were acquaintances of one of the authors. Results will be reported in a future paper. ESM assessments began in June 2025 and will end in December 2025.

As of September 2025, 91 participants have been enrolled for the first wave of ESM. To address recruitment challenges, the time since loss criteria was extended from 3-6 months to within 3 years. The study team also decided to assess the acceptability and feasibility of studying daily life grief reactions via a mobile app using the Reactions to Research Participation Questionnaire adapted for grief-ESM research (Lenferink et al., 2022; Newman et al., 2001). Feasibility will be assessed by reporting compliance and retention rates, and asking participants if their grief prompted professional help in the past two weeks. Presented at the Society of Ambulatory Assessment in Leuven, Belgium, in May 2025, results were not reported due to ongoing data collection. Feasibility and acceptability results will be presented at the Global South Collective Symposium in Ha Noi, Viet Nam, in August 2025. Within-person daily PGD-reaction fluctuations of the first 33 participants will be analyzed and presented at the World Psychiatry Congress in Prague, Czech Republic, in October 2025. Results will be disseminated at international conferences, published in journals, and shared on social media and healthcare websites.

## Discussion

### Significance of expected findings

PGD, despite being a newly introduced mental health condition, has been investigated intensively across different cultures in terms of its progression and comorbidity (Adiukwu et al., 2022; Boelen and Lenferink, 2022; Killikelly et al., 2023; Stelzer et al., 2020). However, little is known about the disorder’s trajectory among Vietnamese people. Therefore, this study poses an effort to investigate the development, progression, and predictive factors of Prolonged Grief Disorder among Vietnamese individuals. Leveraging the advantages of a digital mental health method in ESM, we aim to gather insights into the temporal symptom dynamics, developmental trajectory, and symptom variability of PGD in a low-resource, culturally distinct context.

Given the high risk of grief-related disorders in Viet Nam due to the COVID-19 pandemic, it’s crucial to investigate grief symptoms and their trajectory after losing a loved one. PGD often co-occurs with PTSD and depression (Wen et al., 2023), so understanding the differences between these conditions will provide effective support. Among Taiwanese participants, bidirectional temporal relationships between depressive and PGD symptoms were found in the first year post-loss, and PGD symptoms predicted PTSD symptoms in the second year (Wen et al., 2023). Despite interconnected relationships, yearning and longing are core PGD symptoms distinct from other disorders (Killikelly et al., 2019; Malgaroli et al., 2018). However, the centrality of yearning as a PGD symptom varies across cultures (Mazza et al., 2025). Our study aims to understand the role of cultural context in PGD symptoms. Negative appraisals and loss-related memories are also hypothesized to predict PGD development (Smith & Ehlers, 2021).

An ESM approach in this study investigates PGD symptoms progression, revealing insights into clinical relevance, early detection, risk factors, and intervention periods. Eisma et al. (2022) used the daily diary method to study rumination, worry, and affect in prolonged grief, finding strong interrelations between negative thoughts, affect, and PGD symptoms, suggesting targeted intervention for post-loss adaptation. Lenferink et al. (2024) studied 38 bereaved people in Germany and the Netherlands with an ESM design, finding that grief symptoms fluctuate, challenging theories of a fixed state. Despite ESM’s promising potential, its application in mental health research is scarce. Our study addresses this gap, offering novel insights into daily symptom interactions and predictive factors of PGD in Viet Nam, enriching cross-cultural understandings and promoting innovative methods like ESM.

ESM offers novel insights into symptom variability and interactions, making it a promising tool for developing early detection systems. Fried et al. (2024) proposed an early warning system for depression, incorporating ESM as a key component. Peatzold et al. (2021) identified negative symptom manifestations as predictors of future psychosis outcomes based on ESM data collected over 6 days. ESM-supported early detection can identify critical transitions in mental health deterioration, enabling early intervention before clinical symptoms emerge. Vietnam’s shift towards digitalization and technological integration in mental health aligns with its post-COVID push for digital health transformation. The m-Path’s ease of use and high tailoring make it an opportunity to position app-based early detection tools as a low-cost, scalable approach to identifying individuals at risk before symptoms worsen. Timely, real-world monitoring through this method can bridge care gaps and foster proactive responses in Vietnam’s evolving digital health landscape.

Clinically, ESM-based insights into symptoms, context, contingencies, and daily functioning can also inform momentary self-help modules for everyday life interventions (Mestdagh et al., 2023). Within the app-based ESM environment, clinicians can extend basic assessment with periodic EMI reminders, automated supportive messages, or therapeutic exercises to intervene with the fluctuating nature of grief in everyday life, creating just-in-time adaptive interventions (McDevitt-Murphy et al., 2018; Schueller et al., 2017). Technologically, it advocates for and advances the digitization of mental health in Viet Nam by utilizing app-based ESM methodology, a modern and ecologically valid approach to studying psychological phenomena in daily life. Besides that, as the application of technology in healthcare is growing (Marcolino et al., 2018) and is well-received by stakeholders in Southeast Asia (Pham et al., 2025), this research contributes to the evidence-based support to integrate digital tools in mental health assessment and monitoring, promoting scalable and accessible solutions while setting the foundation for future mental health intervention tools in Vietnam. Furthermore, this research can raise awareness about PGD as a legitimate mental health condition and help destigmatize grief-related psychological distress, particularly when public expression of grief-related distress might not be appropriate (Le et al., 2025).

### Limitations

Despite its innovations, the study is not without limitations. The ESM design, while enhancing ecological validity, relies heavily on participants’ consistent and honest engagement, where certain concerns, such as haphazard responses and contextual biases due to daily ESM prompt scheduling, are raised (Trang et al., 2022; Doherty et al., 2020). Moreover, prolonged exposure to similarly themed messages throughout the ESM data collection can result in low compliance due to message fatigue (Kim & So, 2018). Additionally, our intended sample may also not be representative of the full cultural and socioeconomic diversity within Viet Nam, limiting generalizability. For instance, digital access and technological literacy may influence who is able or willing to participate in an ESM-based study. Older adults or individuals in rural areas may have limited access to smartphones or feel less comfortable using app-based platforms, which could introduce participation bias and affect compliance. The potential scale-up of ESM for digital screening and diagnosing of mental health conditions faces certain challenges since mental healthcare in Viet Nam is at an early stage of development, largely due to limited resources (e.g., a shortage of professionals, inadequate facilities, and low levels of mental health literacy and awareness) (Minas et al., 2017). Lastly, the observational nature of the study constrains causal inferences from the findings.

Being the first study on PGD in Vietnam, we expect to lay the foundations for understanding the disorder in the Vietnamese context. Future research should explore diagnostic applicability, including clinical thresholds, culture-specific manifestations, comorbidity, and epidemiology. Research advancing mental health digitalization in Vietnam, such as with various data types or digital interventions, is also recommended. Studies exploring PGD symptom variability, developmental trajectory, and disorder expression across cultures are suggested to provide nuanced cross-cultural insights.

## Conclusion

This study aims to understand PGD in the Vietnamese context, where grief-related mental health conditions are under-researched. Using ESM, we’ll capture PGD’s real-time dynamics, developmental trajectory, and predictive factors, providing insights into its cultural expression, symptom variability, and progression. As the first longitudinal study to gather and analyze large-scale, systematic data on bereavement in Viet Nam, we’ll expand cross-cultural scientific knowledge and advance mental health digitalization. We anticipate contributing to early detection tools, clinical interventions, and public awareness efforts to destigmatize grief-related psychological distress. This study lays a strong foundation for PGD research in Viet Nam and contributes to the country’s focus on digital health. It advocates for more nuanced, culturally informed, and technologically integrated approaches to bereavement care in Viet Nam and other low-resource settings.

## Data Availability

All data produced in the present study are available upon reasonable request to the authors

